# Optimal timing for social distancing during an epidemic

**DOI:** 10.1101/2020.03.30.20048132

**Authors:** Oscar Patterson-Lomba

## Abstract

Social distancing is an effective way to contain the spread of a contagious disease, particularly when facing a novel pathogen and no pharmacological interventions are available. In such cases, conventional wisdom suggests that social distancing measures should be introduced as soon as possible after the beginning of an outbreak to more effectively mitigate the spread of the disease. Using a simple epidemiological model we show that, however, there is in fact an *optimal* time to initiate a temporal social distancing intervention if the goal is to reduce the final epidemic size or “flatten” the epidemic curve. The optimal timing depends strongly on the effective reproduction number (*R*_0_) of the disease, such that as the *R*_0_ increases, the optimal time decreases non-linearly. Additionally, if pharmacological interventions (e.g., a vaccine) become available at some point during the epidemic, the sooner these interventions become available the sooner social distancing should be initiated to maximize its effectiveness. Although based on a simple model, we hope that these insights inspire further investigations within the context of more complex and data-driven epidemiological models, and can ultimately help decision makers to improve temporal social distancing policies to mitigate the spread of epidemics.

## Introduction

As the SARS-CoV-2 virus sweeps through the globe, the concept of “social distancing” has become mainstream across news outlets [14, 22, 21, 18], government announcements [2, 1] and research publications [16, 7, 25, 26, 5, 12, 4]. Social distancing is a very effective way to slow down the swift advance of the COVID-19 pandemic, especially given the current lack of effective pharmaceutical interventions, and our limited ability to identify, let alone track, infectious individuals, particularly asymptomatic ones.

Aiming to reduce instances of person to person transmission, social distancing is a measure that is customarily put into action to decrease the number of contacts among individuals [24]. Examples of social distancing include closing of schools, non-essential workplace closures, and avoiding large gatherings (i.e. public transport, concerts, religious gatherings) in which a large number of individuals are in close contact and facilitating the contagion process [15]. In most cases, due to socio-economic and logistic reasons [13], social distancing measures can only be in effect for a finite period of time (e.g., 2 weeks, 1 month, 2 months) that is typically much shorter than the full duration of the epidemic.

Conventional wisdom suggests that the sooner social distancing measures are implemented, the higher the chances of curtailing the spread of the virus. Prompt and strict social distancing measures can help “flatten the epidemic” (another fashionable meme at the moment), which in turn will help prevent overwhelming the health system, while also buying us time until effective antivirals and vaccines can be deployed on a mass scale [19]. Implementing swift social distancing measures is one of the most effective ways to stop the spread of the virus, and when the facing a novel and very transmissible virus like COVID-19, social distancing is our first and most effective line of defense. However, is it true that the *sooner* a temporal social distancing measured is introduced the better health outcomes we will get? The answer to this question would be a resounding “yes” if such measures were to last for extended periods of time (i.e., until the epidemic is over or almost over). However, social distancing interventions are seldom extended long enough to drive the epidemic to extinction on its own. In fact, it is known that new epidemic waves can arise after the social distancing measures are lifted if the pool of susceptibles is large enough, as current models suggest [4, 15, 11], and as it has historically happened in several occasions, including during the 1918 Spanish influenza pandemic [6], the 2003 SARS epidemic in Canada [8], and the 2009 H1N1 influenza epidemic in Mexico [10].

We argue that how soon temporal social distancing should start depends on what are the health outcomes we are optimizing for, and on when other effective interventions (such as antivirals or vaccines) will be available following the social distancing period. If the goal is to reduce the final number of infected cases, or reduce or delay the peak of the epidemic (i.e., flatten the epidemic curve), we show the existence of an *optimal* timing to initiate the social distancing period.

The goal of this paper is to study how to efficiently *time* temporal social distancing measures, and investigate how the optimal time to implement a social distancing intervention depends on i) disease transmissibility, ii) the length of the social distancing, iii) its effectiveness in reducing transmissibility, and iv) when will other (pharmacological) interventions will be available. We define” optimal” based on three different epidemiologically relevant criteria: optimal timing of social distancing 1) to minimize the final size of the epidemic; 2) to maximize the delay in the peak of the infection incidence curve of the epidemic; and 3) to minimize the peak of the infection incidence curve of the epidemic. The first criteria aims to minimize the impact of the epidemic in terms of overall infections, while the other two refer to the flattening of the epidemic as to not overburden the health system in place to care for infected individuals.

## Methods

### The SIR model with social distancing

The idea of an optimal social distancing timing is investigated in the context of a classic SIR type model [3]. Individuals are classified based on their infectious status as susceptible (*S*), infected (*I*) and recovered (*R*). The force of infection at time *t* is given by *β*(*t*)*I*(*t*), where *β*(*t*) is the transmission probability. This term is the product of the number of contacts per capita per time unit, *c*(*t*), and the probability of transmission given a contact, *p*; that is *β*(*t*) = *c*(*t*)*p*. The social distancing measure affects the *c*(*t*) parameter, such that when social distancing is in effect *c*(*t*) is reduced. The corresponding reduction in *β*(*t*) due to a temporary social distancing measure can be modeled as a “rectangular well” function, as shown in Figure (1). From the start of the epidemic until time *t*_0_ (the time at which social distancing is initiated), the transmission rate is *β*. From time *t*_0_ to *t*_0_ + *T* a social distancing measured is implemented such that the transmission rate is reduced to *β*_*r*_ = *rβ*, with 0 *≤ r ≤* 1, and *T* being the length of the period when social distancing measures are in place. Then, after time *t*_0_ + *T* (when social distancing ends), the transmission rate comes back up to *β*, until time *t*_*I*_, at which point a fully effective intervention (e.g., vaccine) is applied such that transmission is completed halted; that is, the effective reproduction number is zero for *t* > *t*_*I*_ (for simplicity we model such scenario by assuming that *β* = 0).

**Figure 1:**
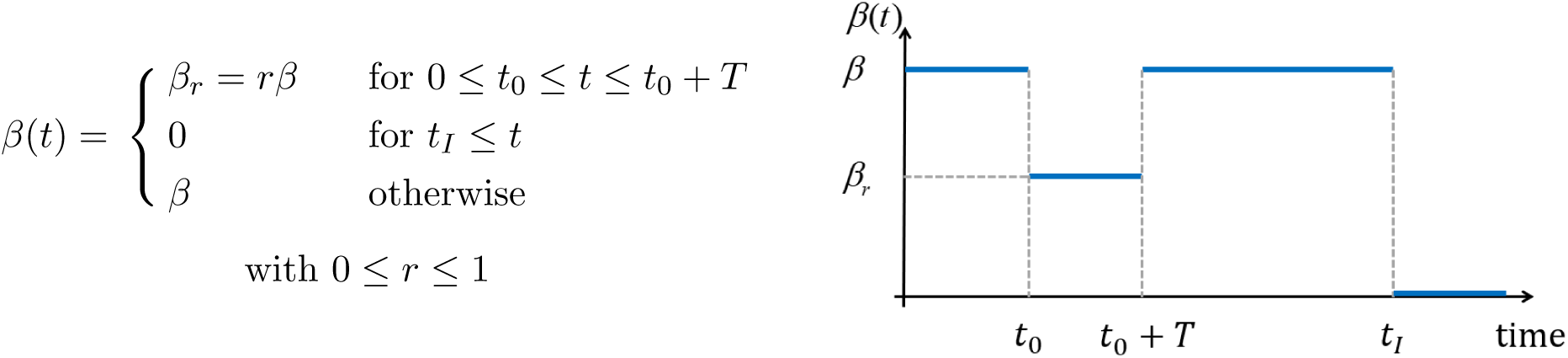
Time-dependent transmission rate. From time *t*_0_ to *t*_0_ + *T* a social distancing measured is implemented such that the transmission rate is reduced by a factor *r*. At time *t*_*I*_ an intervention (e.g., vaccine) is applied such that *β* = 0.

The system of nonlinear differential equations describing the disease spread dynamics is given by

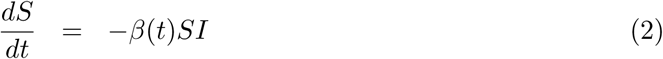

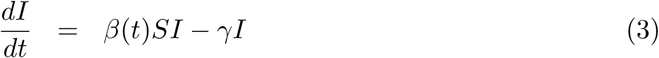

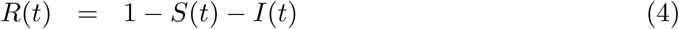

where, with no loss of generality, the size of the population is assumed to be 1 and to remain constant (assuming no births and death processes). The parameter *γ* is the recovery rate. The ODE system in (2)-(3) has a basic reproductive number given by *R*_0_ = *β*(*t*)*/γ*.

In Appendix 1 we derived the expression for the final epidemic size (i.e., the fraction of infected individuals at the end of the epidemic) in the context of temporal social distancing measure. The resulting expression implicitly relates the final epidemic size, *y*, and *t*_0_ in a transcendental equation. Thus, *y* cannot be isolated as to allow a close form solution to the optimal *t*_0_ that minimizes *y*.

### Numerical simulations setup

We use numerical simulations of the model in (2)-(3) with the goal to find the optimal *t*_0_ for reducing the final epidemic size as a function of *R*_0_, the length of the social distancing intervention (*T*) and its effectiveness in reducing disease transmissibility (*r*). The model was simulated for 5 years over a wide range of values for *R*_0_, *T* and *r* (see Appendix 2 for details). For the first set of simulations, we assumed that *t*_*I*_ = *∞* (i.e., no pharmacological intervention becomes available during the epidemic).

For each combination of *R*_0_, *T* and *r*, the value of *t*_0_ corresponding to the minimal final size was obtained. Moreover, to quantify the effect of the optimal social distancing timing, the reduction in final size corresponding to optimal timing (as compared to a scenario with no social distancing) was also computed.

We also use numerical simulations to find the optimal *t*_0_ for, first, delaying the peak of the epidemic curve, and second, for reducing the peak (i.e., the maximum prevalence of cases), as a function of *R*_0_, *T* and *r*. To quantify the effect of these optimal social distancing timings, the delay in the peak and the reduction in the prevalence peak corresponding to optimal timing (as compared to a scenario with no social distancing) were also computed. For the next set of simulations we relax the assumption about *t*_*I*_, and instead assume that *t*_*I*_ ∈ (60 − 730) days. For each value of *t*_*I*_, it is assumed that *t*_0_ + *T < t*_*I*_. For these simulations, we set *r* = 0.55 and *T* = 30 days for simplicity. The optimal timing for social distancing is then computed for each value of *R*_0_ and *t*_*I*_.

## Results

### Preliminary insights

Figure (2) show simulations of the model in (2)-(3) with different temporal social distancing scenarios. In particular, we explore the impact of different times at which the social distancing measures are implemented (*t*_0_) in the disease dynamics and the final epidemic size. From Figure (2) (left) we see that if the social distancing is implemented *too soon* (*t*_0_ = 50 days), then at the end of the social distancing measure (after 30 days) there are still some infected in the population, and the pool of susceptible is large enough (low population-level immunity) for the disease to take off again and still infect a large portion of the population. Conversely, if social distancing is introduced *too late* (*t*_0_ = 100 days) by the time the epidemic has already largely spread in the population, then social distancing measures less effective in reducing the final epidemic size. In both these cases, social distancing measures were not applied in an optimal manner, time wise. However, if social distancing is initiated at *t*_0_ = 80 then the resulting epidemic is more effectively flattened. These observations are crystalized in Figure (2) (right), which suggests that there is in fact an optimal time, 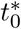, to introduce the social distancing so that the final epidemic size is minimized. Moreover, this figure suggests that the *t*_0_ that minimizes the final epidemic size decreases as *R*_0_ increases. That is, the more transmissible the disease is, the sooner social distancing measures should be initiated.

**Figure 2:**
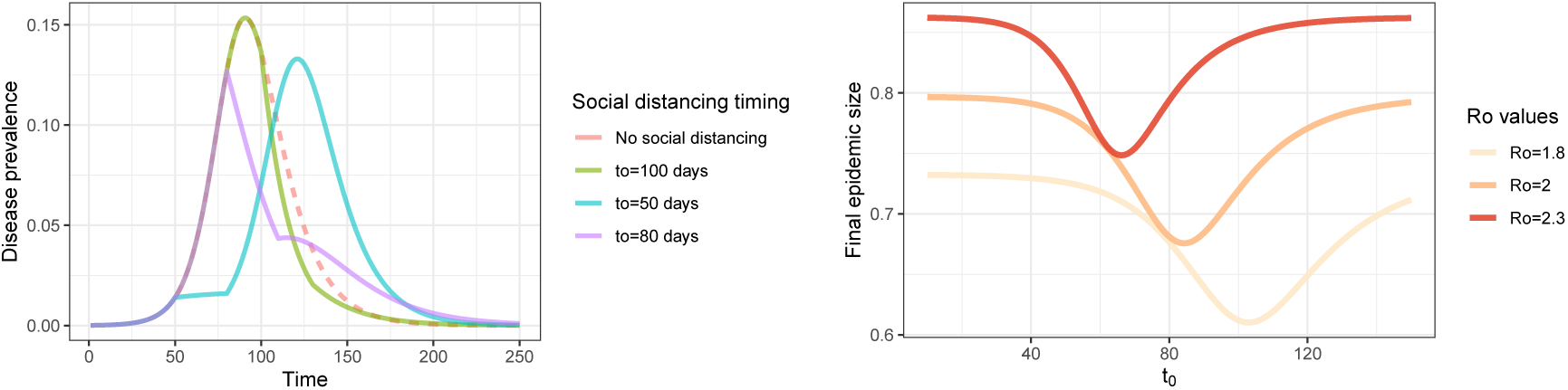
(left) Disease prevalence for different *t*_0_ values and no social distancing (dashed line) with *R*_0_ = 2. (right) Final epidemic size for different values of *t*_0_ for three different values of *R*_0_. For all simulations, *γ* = 1*/*10, *T* = 30 days, *r* = 0.55, *I*(0) = 0.01%.

### Simulations for the optimal *t*_0_: final size

Building on the findings from the previous section, in what follows we conduct a more comprehensive analysis of how the optimal timing of social distancing depends on *R*_0_, *r* and *T* when the objective is to minimize the final epidemic size. Figure (3) indicates that *R*_0_ is the key factor in determining *when* to initiate social distancing, with *r* and *T* having virtually no effect. As *R*_0_ increases, the sooner social distancing should start (see Figure (6)). Importantly, the optimal timing is never immediately after the beginning of the epidemic, unless *R*_0_ is extremely large.

**Figure 3:**
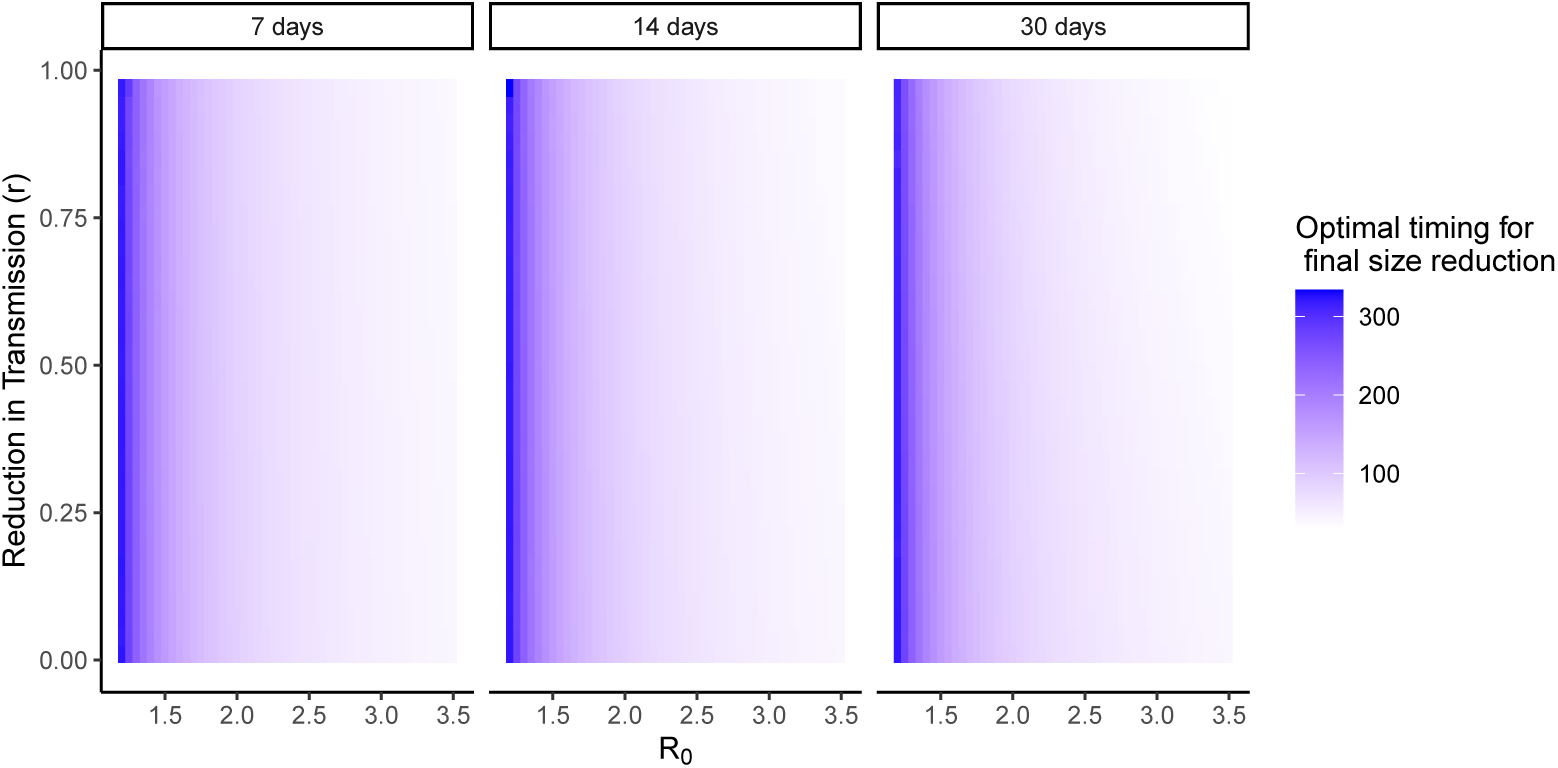
Optimal *t*_0_ to initiate social distancing when the objective is to minimize the final epidemic size, as a function of *R*_0_ (x axis), reduction in transmissibility during social distancing *r* (y axis), and length of social distancing intervention (7, 14 and 30 days). As *R*_0_ increases, the smaller the optimal time to initiate social distancing. For all simulations *γ* = 1*/*10.

The effects of *r* and *T* are important in terms of the extent to which an optimally timed social distancing can reduce the final epidemic size (see Figure (8) in Appendix 3), with longer *T* and smaller *r* yielding greater reductions (up to 35% reductions in final size with the parameter ranges explored). Interestingly, the lower the *R*_0_ the greater the reduction in final size.

### Simulations for the optimal *t*_0_: flattening the epidemic

#### Optimal *t*_0_ for peak delay

Figure (4) also indicates that *R*_0_ is the key factor in determining the optimal time to initiate social distancing if the goal is to delay the epidemic peak. Again, as *R*_0_ increases, the sooner social distancing should start (see Figure (6)). Longer *T* and smaller *r* yield greater delays (up to 250 days), as expected (see Figure (9) in Appendix 3). Finally, the lower the *R*_0_ the greater the delay in peak timing.

**Figure 4:**
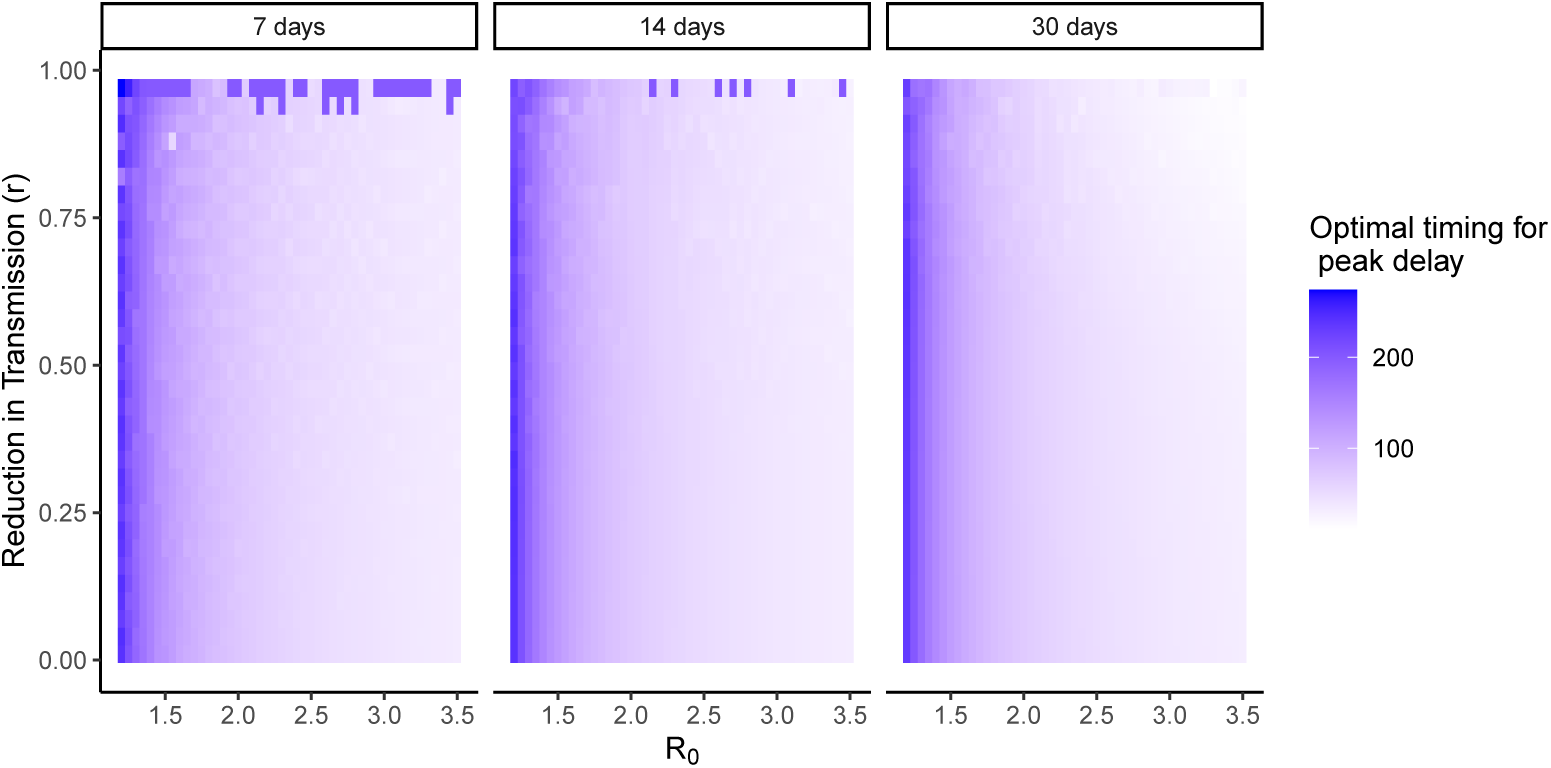
Optimal *t*_0_ to initiate social distancing if the goal is to delay the epidemic peak, as a function of *R*_0_ (x axis), reduction in transmissibility during social distancing *r* (y axis), and length of social distancing intervention (7, 14 and 30 days). As *R*_0_ increases, the smaller the optimal time to initiate social distancing. For all simulations *γ* = 1*/*10.

#### Optimal *t*_0_ for peak reduction

Figure (5) again indicates that *R*_0_ is the main factor determining the optimal time to initiate social distancing if the goal is to reduce the epidemic peak. Once more, as *R*_0_ increases, the sooner social distancing should start (see Figure (6)). Longer *T* and smaller *r* yield greater peak reductions (up to 60% reductions), (see Figure (10) in Appendix 3). Finally, the higher the *R*_0_ the greater the reduction in peak prevalence.

**Figure 5:**
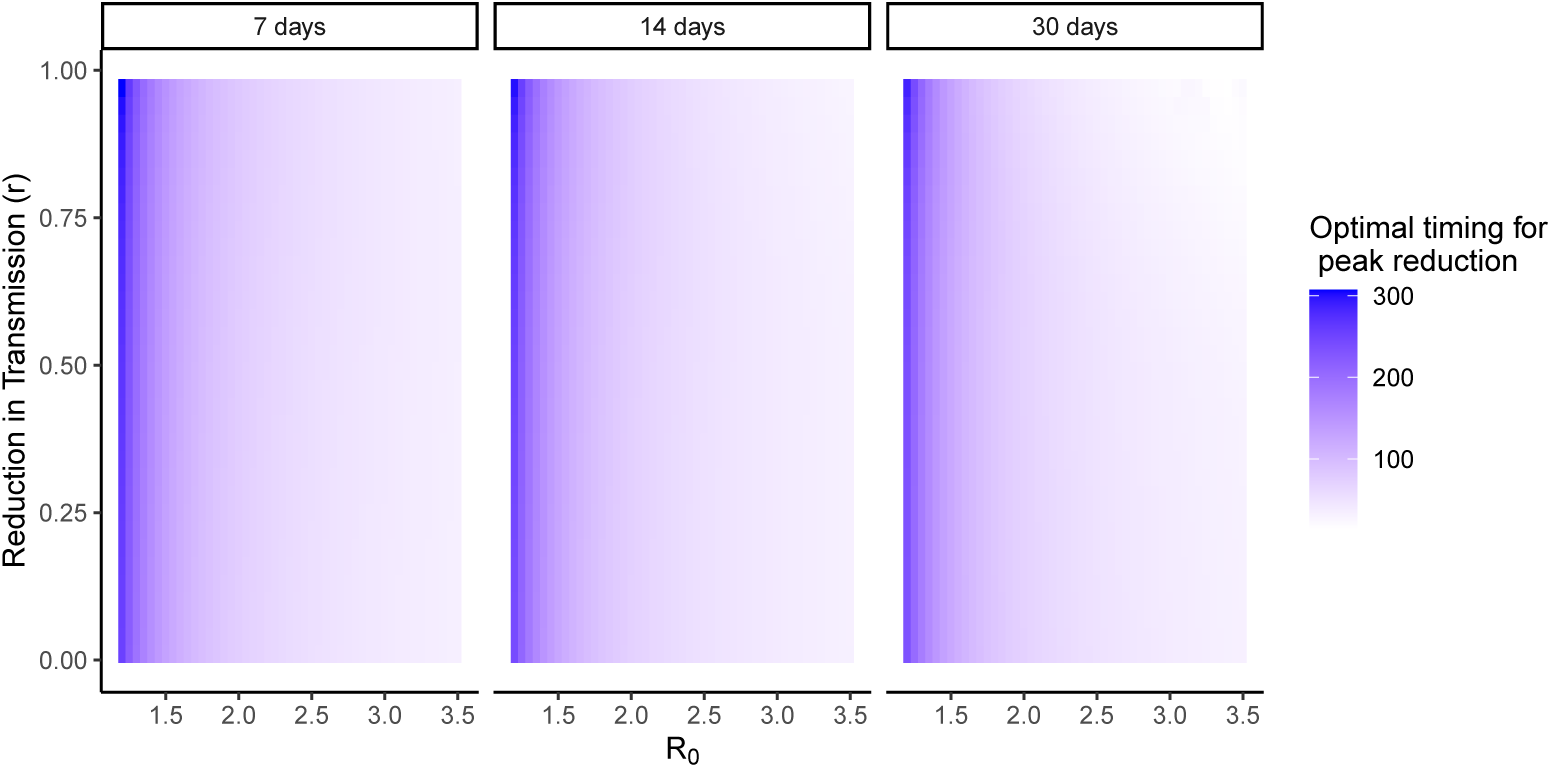
Optimal *t*_0_ to initiate social distancing if the goal is to reduce the epidemic peak, as a function of *R*_0_ (x axis), reduction in transmissibility during social distancing *r* (y axis), and length of social distancing intervention (7, 14 and 30 days). As *R*_0_ increases, the smaller the optimal time to initiate social distancing. For all simulations *γ* = 1*/*10.

Figure (6) shows that the optimal timing for social distancing decreases with *R*_0_ in a quasi-exponential way, regardless of the epidemic containing strategy being employed. Interestingly, if the objective is to minimize final size, then the social distancing should start later compared to a social distancing strategy aimed at flattening the epidemic (either reducing or delaying the peak), with the differences decreasing as *R*_0_ increases. For example, for an epidemic with *R*_0_ = 1.5 (i.e., typical of a flu epidemic), the optimal social distance should be initiated 150 days after the start of the epidemic^1^ if the goal is to minimize the final epidemic size, whereas if the goal is to delay or reduce the peak as much as possible, social distancing should start at about 120 days.

**Figure 6:**
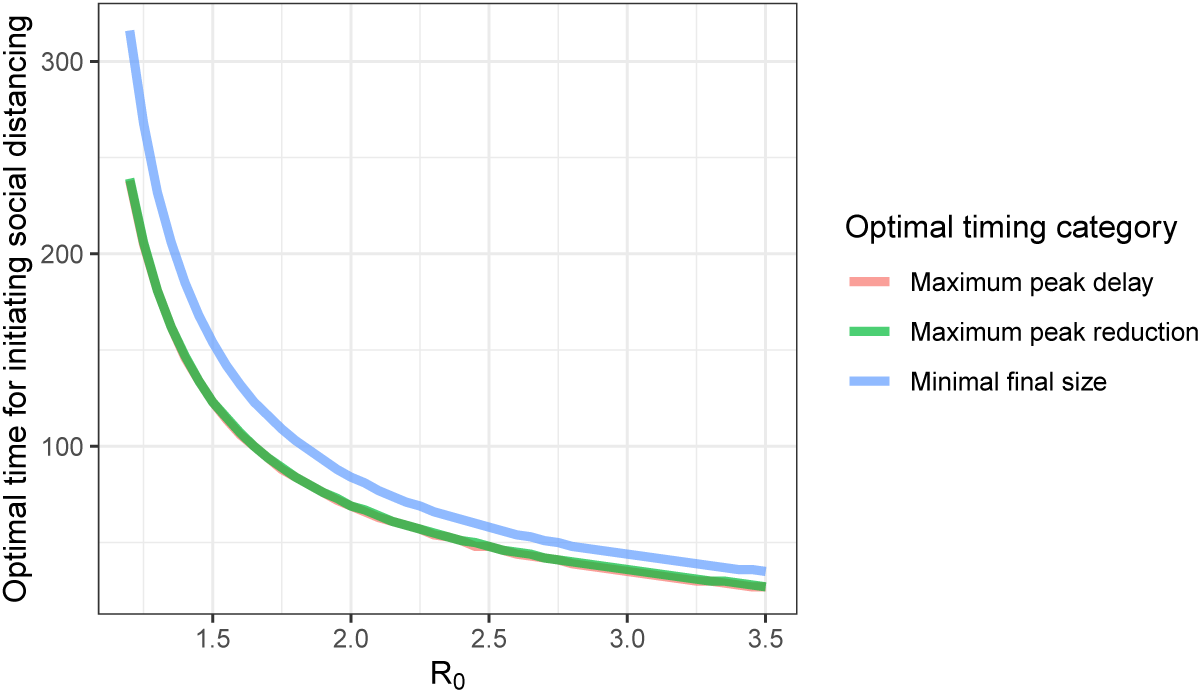
Optimal timing for social distancing vs *R*_0_ for the three different types of optimal timing investigated, and corresponding to *T* = 30 days and *r* = 0.55 (very similar results are obtained for other values of *T* and *r*). Note that the curves for “maximum peak delay” and “maximum peak reduction” are overlaid.

### Simulations for the optimal *t*_0_ when an effective intervention is available in the future

So far, the model has assumed that the only available intervention to curtail the spread of the disease is social distancing, and that other pharmacological interventions are not available at any point; or put differently, *t*_*I*_ = *∞*. This assumption is most often wrong, as treatment options (e.g., vaccines, antivirals) typically become available at some point during the course of the epidemic. Here we explore a model with finite values of *t*_*I*_, and see how the optimal social distancing time varies with *t*_*I*_ and *R*_0_. Figure (7) (left) shows that as *t*_*I*_ decreases so does the optimal social distancing time if the goal is to reduce the final size (similar results are obtained if the goal is to flatten the epidemic curve, see Appendix 4). If the epidemic is only moderately transmissible (e.g., 1 *< R*_0_ *<* 2) and pharmacological interventions are available relatively quickly (e.g., *t*_*I*_ *<* 200 days), the optimal social distancing should be initiated almost immediately after the start of the epidemic. Intuitively, this result makes sense; if effective interventions become available soon after the start of the epidemic, a large second wave post social distancing is less likely because the intervention would prevent it.

**Figure 7:**
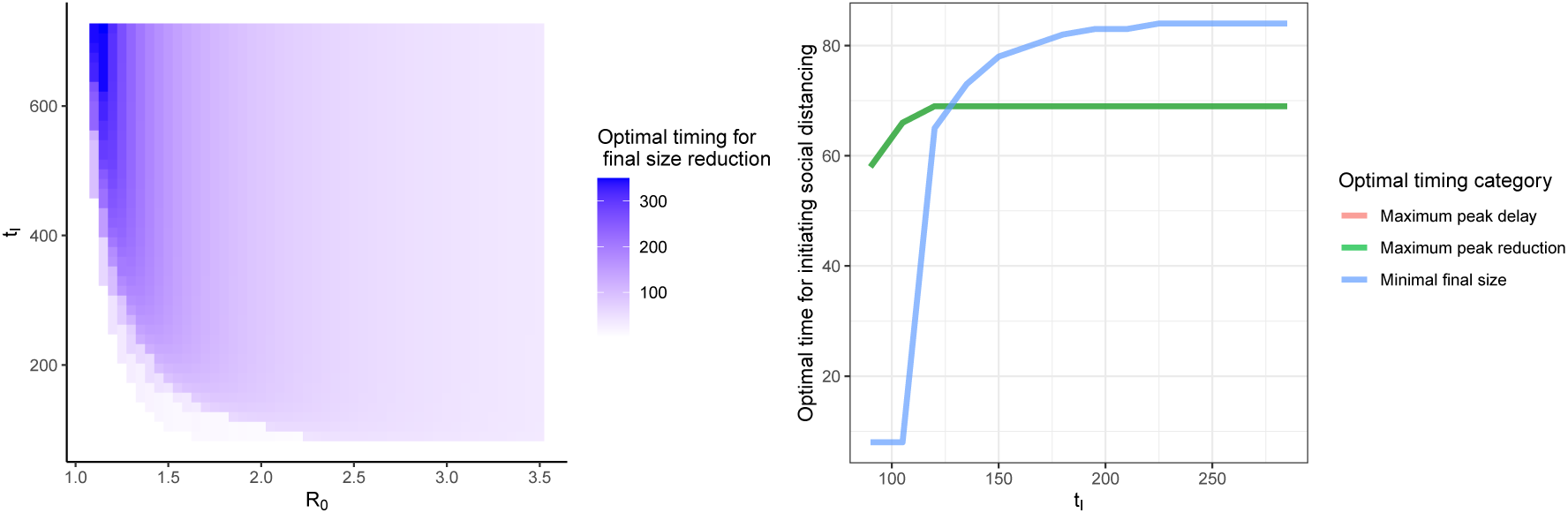
(left) Optimal *t*_0_ to initiate social distancing to minimize final epidemic size as a function of *R*_0_ (x axis), and time to pharmacological intervention (*t*_*I*_) (y axis). (right) Optimal timing for social distancing vs *t*_*I*_ for the three different types of optimal timing investigated, and corresponding to *R*_0_ = 2. With *γ* = 1*/*10, *T* = 30, *r* = 0.55, *I*(0) = 0.01%. Note that the curves for “maximum peak delay” and “maximum peak reduction” are overlaid.

Figure (7) (right) shows that the optimal timing for social distancing decreases sharply (particulalrly if the goal is to reduce the final size) as *t*_*I*_ decreases, regardless of the epidemic containing strategy being employed. Note that if *t*_*I*_ is large enough (i.e., the intervention comes long after the peak of the epidemic with no containing measures [*∼* 90 days, see Figure (2) (left)]), then the optimal time stops depending on the value of *t*_*I*_. In the previous section we noted that if the objective is to minimize final size, then the social distancing should start later compared to a social distancing strategy aimed at flattening the epidemic. This is indeed the case for large *t*_*I*_; however, for low values of *t*_*I*_, the optimal time to minimize final size is shorter compared to a strategy aimed at flattening the epidemic.

## Discussion

We investigated the idea of optimally timing the start of temporary social distancing measures to more effectively hinder the spread of an epidemic. We find that, for a given disease transmissibility, and social distancing transmission-reduction effectiveness and length, there exists an optimal time to initiate the social distancing intervention. Regardless of the criteria used to define “optimum” timing, let it be final epidemic size reduction, epidemic peak delay or epidemic peak reduction, the timing is closely related to *R*_0_, and not so much with *T* and *r*. Moreover, we find that the optimal time follows an approximate exponential decay relationship with *R*_0_, such that the higher the *R*_0_ the sooner social distancing measures should be initiated.

The factors *r* and *T* determine the extent to which optimally timed social distancing reduces the final epidemic size or flattens the epidemic curve. Larger reductions in transmissibility and social distancing of longer durations lead, as expected, to grater reductions in final size and more flattening of the epidemic. Interestingly, the lower the *R*_0_ the greater the potential reductions of optimally timed social distancing measures in final size and delay in peak prevalence, whereas the higher the *R*_0_ the greater the reduction in peak prevalence.

If pharmacological interventions, such as effective treatments and/or vaccines, become available at scale, the time point at which they do so has also important implications for the optimal time for social distancing. The sooner pharmacological interventions become available, the sooner social distancing should be introduced, particularly if the pharmacological interventions can be deployed around or right after the time the epidemic would peak in the absence of any sort of containing intervention.

It is important to remark that these results stem from very idealized assumptions about the course of the epidemic. The inclusion of transmission or contact level heterogeneities within the population [19] (e.g., age structure) may play a significant role in the quantitative description of the results in this paper, as would the inclusion of seasonality effects. Additionally, since we are using a deterministic mean-field approach, these analyses do not account for the effects of stochastic fluctuations (which could lead to epidemic extinction if the number of infected is low enough during the social distancing phase) and as a consequence, the findings herein apply more closely to very large populations (i.e., in the thermodynamic limit) where stochastic fluctuations are less relevant. Similarly, the model assumes homogeneous mixing among individuals, thus the contact network structure of the population is not accounted for; as a result this model cannot realistically predict the epidemic-hampering effects that social distancing measures (e.g., limiting the size of crowds) have in the context of contact structured populations [20].

An important assumption in this exercise of finding an optimal time for initiating social distancing, is that individuals that are infected or immune are all perfectly ascertained and tracked (i.e., no underreporting of cases) such that decision makers have perfect knowledge of the *R*_0_ and the *effective* reproduction number (via estimation from epidemic data), as well as the number of new cases at any given point. Early in an epidemic, particularly when in the presence of a novel pathogen like SARS-CoV-2, there is typically a large amount of uncertainty around most epidemiological parameters, especially *R*_0_ [9, 23]. This uncertainty is the result of several factors, including lack of reliable data on the true number of cases, differences in the formulations and assumptions underlying the statistical models used to estimate *R*_0_, and the geographic heterogeneity of the disease spreading patterns. This uncertainty around the value of *R*_0_ should also be carried over to the estimate of the optimal time for initiating social distancing. Hence, assuming the true *R*_0_ lies within a range (e.g., 95% confidence intervals), the estimation of the optimal *t*_0_ should account for this uncertainty. Moreover, if a (Bayesian posterior) distribution is available for *R*_0_, then one could also get a distribution for the optimal *t*_0_, which would in turn allow to compute the probability that the optimal *t*_0_ is within a given range, or lower/higher than a given time of interest (e.g., two weeks from now). Then, it would be up to the decision makers to determine what level of uncertainty is considered acceptable in order to proceed with a given social distancing strategy. Of note, if the level of uncertainty on *R*_0_ is very large and a decision needs to be made promptly, it is arguably better to initiate social distancing sooner, as opposed to later, than the optimal *t*_0_ inferred from the available data; therefore, a conservative approach could be to assume *R*_0_ is equal to the upper bound of its %95 confidence interval, and compute the corresponding (shorter) optimal *t*_0_.

The findings herein serve to show the important “proof of concept” of an optimal time to initiate social distancing, which arises as an interesting and potentially useful feature of a simple epidemiological model. Our hope is that these insights inspire other researchers to investigate the existence of optimal (or just more effective) times to implement social distancing measures within the context of more complex and data-driven epidemiological models, as to better serve decision makers developing strategic policies to mitigate the extent of current and future epidemics.

## Data Availability

No empirical data was used in this manuscript. Results are based on analytical derivations and numerical simulations.

## Acknowledgments

We would like to thank Andres Gomez-Lievano for helpful comments and suggestions on a previous draft of this manuscript.

## Appendix 1: Derivation of final size expression

Here we find an analytical expression for the final epidemic size (i.e., the fraction of infected individuals at the end of the epidemic) in the context of temporal social distancing measure. For this derivation, we assume that *t*_*I*_ = ∞. In principle, this expression should hold a connection between the final epidemic size and *t*_0_. The ODE system in (2)-(3) has a basic reproductive number given by *R*_0_ = *β*(*t*)*/γ* (with *t* ≈ 0). With no social distancing being implemented, that is *β*(*t*) = *β*, the *final size relation* for the system (2)-(3) is:

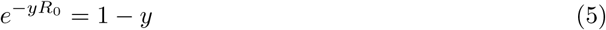

Lets now compute the same final size relation for when the transmission rate corresponds to the piecewise function shown above in Figure(1), i.e., it is reduced for a period of time *T* Assume that at the beginning of the epidemic the population was divided into susceptible (*S*_0_) and infected (*I*_0_), such that *N* = *I*_0_ + *S*_0_. We proceed to compute the final size as follows:

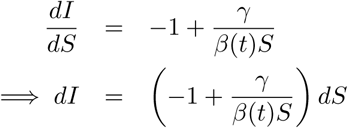

Integrating with respect to time until time *τ* taking *τ* > *t*_0_ + *T* we get

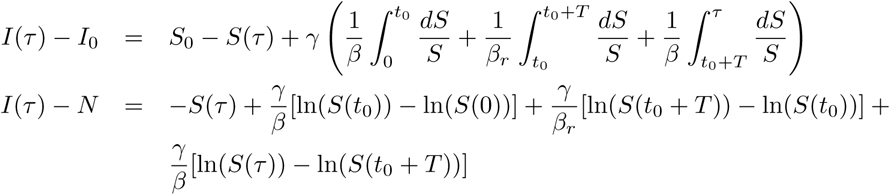

Assuming that *S*(0) ≈ 1 and taking the limit *τ* → ∞, and after a number of algebraic manipulations, while defining *f* as the final size proportion of infected (attack rate) with SD, we obtain

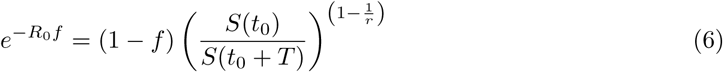

where *f* is the epidemic final size (the proportion of individuals that got infected during the outbreak). Thus, when the transmission rate corresponds to the piecewise function shown in Figure (1), the final size expression is instead given by:

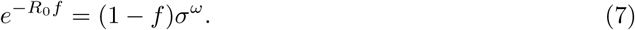

where

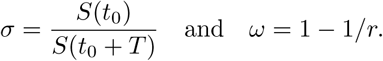

To check the soundness of this expression, note that if *r* = 1 (social distancing was completely ineffective in reducing transmission) then *ω* = 0, and we obtain the classical expression for the final size in Equation (5). Another similar instance is given by *σ* = 1 which corresponds to *T* = 0.

Equation (7) implicitly relates *f* and *t*_0_. The functional form of *f* (*t*_0_) would serve to derive a condition on *t*_0_ that minimizes *f* that is, find 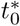 such that min 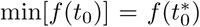. However, such transcendental equation does not allow us to isolate *f*

## Appendix 2: Numerical simulations setup

For each value of *t*_0_ ∈ (0 − 350), *R*_0_ ∈ (1.2 − 3.5), *r* ∈ (0.01− 0.99) and *T* ∈ {7, 14 and 30 days}, the model in system in (2)-(3) was simulated in R[17] for 1825 days (5 years). For all simulations *S*(0) = .9999, *I*(0) = 0.001, and *γ* = 1*/*10.

For each combination of *R*_0_, *T* and *r*, the value of *t*_0_ corresponding to 1) the minimal final size at 5 years 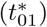, 2) the maximum delay in incidence peak time 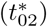, and 3) the minimum incidence peak 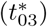 was obtained. To quantify the effect of these three versions of optimal social distancing, a base model with no social distancing was also simulated for each combination of *R*_0_, *T* and *r*. The final size reduction assuming optimal SD timing was computed as:

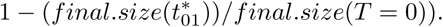

The incidence peak delay assuming optimal SD timing was computed as:

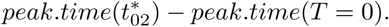

The incidence peak reduction assuming optimal SD timing was computed as:

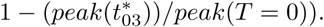

## Appendix 3: Effect of optimal *t*_0_ in reducing final epidemic size and flattening the epidemic

Figure (8) suggests, not surprisingly, that the effects of *r* and *T* are important in terms of the extent to which an optimally timed social distancing can reduce the final epidemic size, with longer *T* and smaller *r* yielding greater reductions (up to 35% reductions in final size with the parameter ranges explored). Interestingly, the lower the *R*_0_ the greater the reduction in final size.

**Figure 8:**
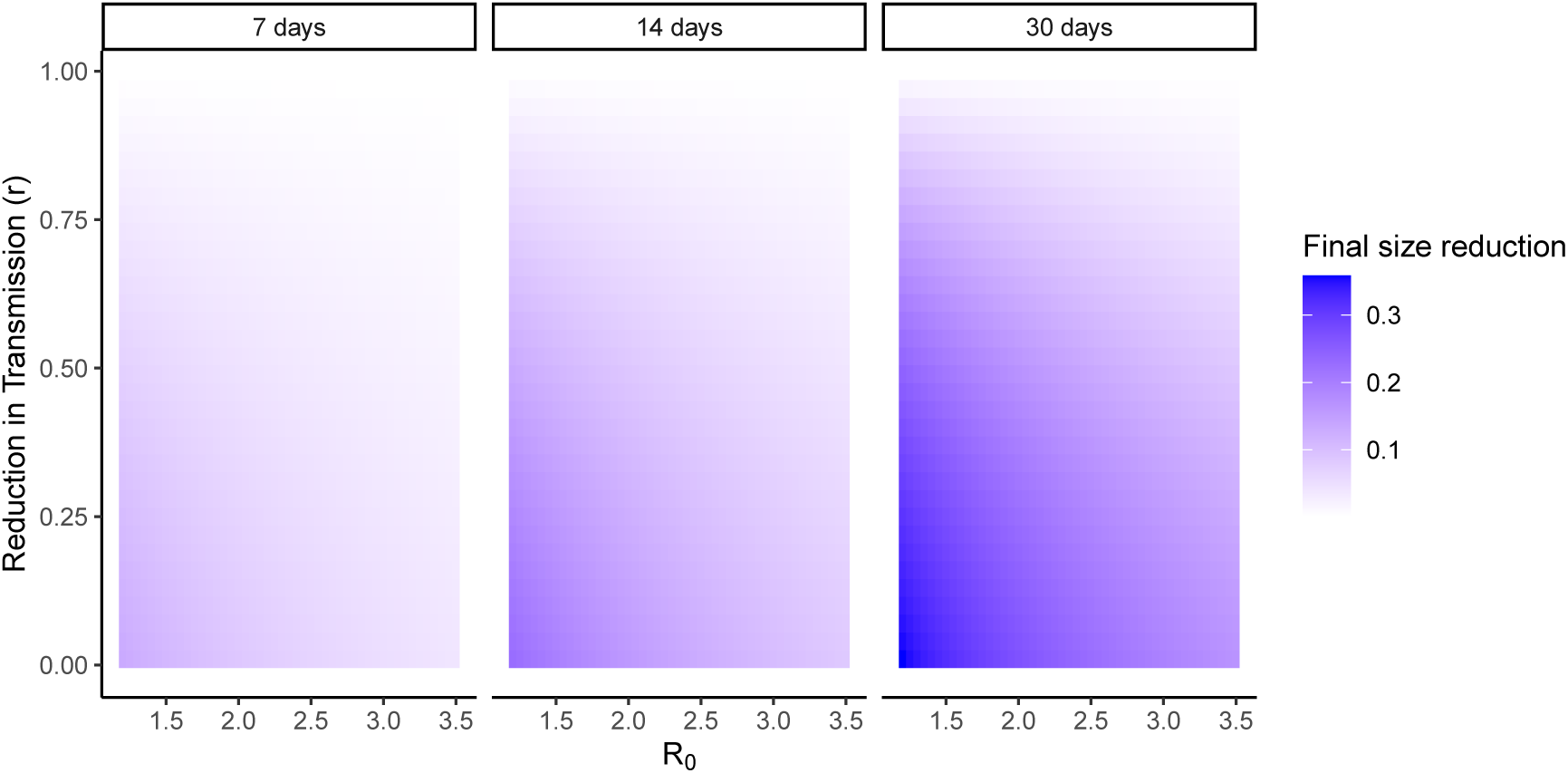
Reduction in final size corresponding to optimal *t*_0_ to initiate social distancing as a function of *R*_0_ (x axis), reduction in transmissibility during social distancing *r* (y axis), and length of social distancing intervention (7, 14 and 30 days). For all simulations *γ* = 1*/*10.

Figure (9) indicates that longer *T* and smaller *r* yield greater delays (up to 250 days), as expected. Finally, the lower the *R*_0_ the greater the delay in peak timing.

**Figure 9:**
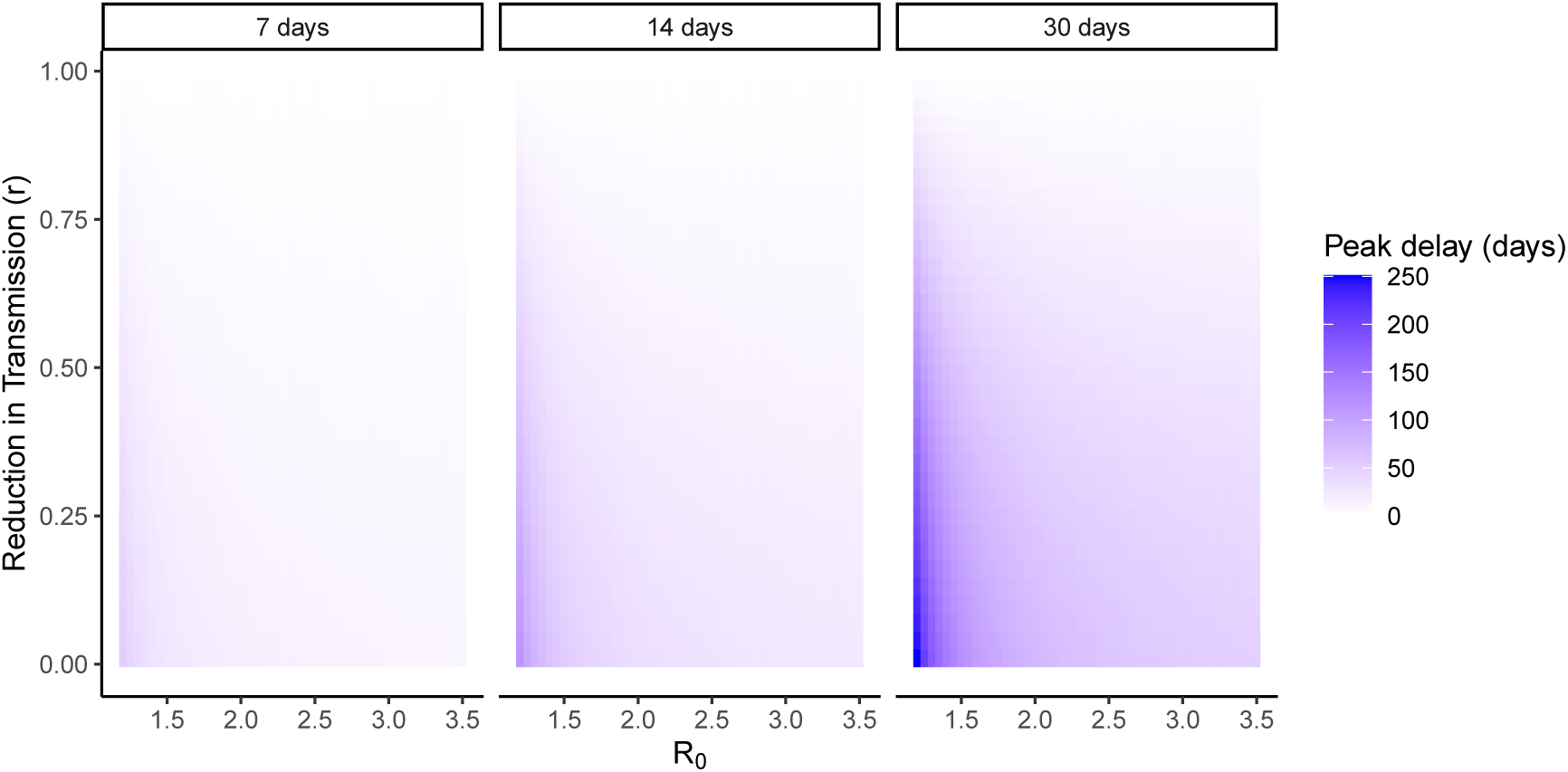
Delay in peak time corresponding to optimal *t*_0_ to initiate social distancing as a function of *R*_0_ (x axis), reduction in transmissibility during social distancing *r* (y axis), and length of social distancing intervention (7, 14 and 30 days). For all simulations *γ* = 1*/*10.

Figure (10) indicates that longer *T* and smaller *r* yield greater peak reductions (up to 60% reductions), as expected. Finally, the higher the *R*_0_ the greater the reduction in peak prevalence.

**Figure 10:**
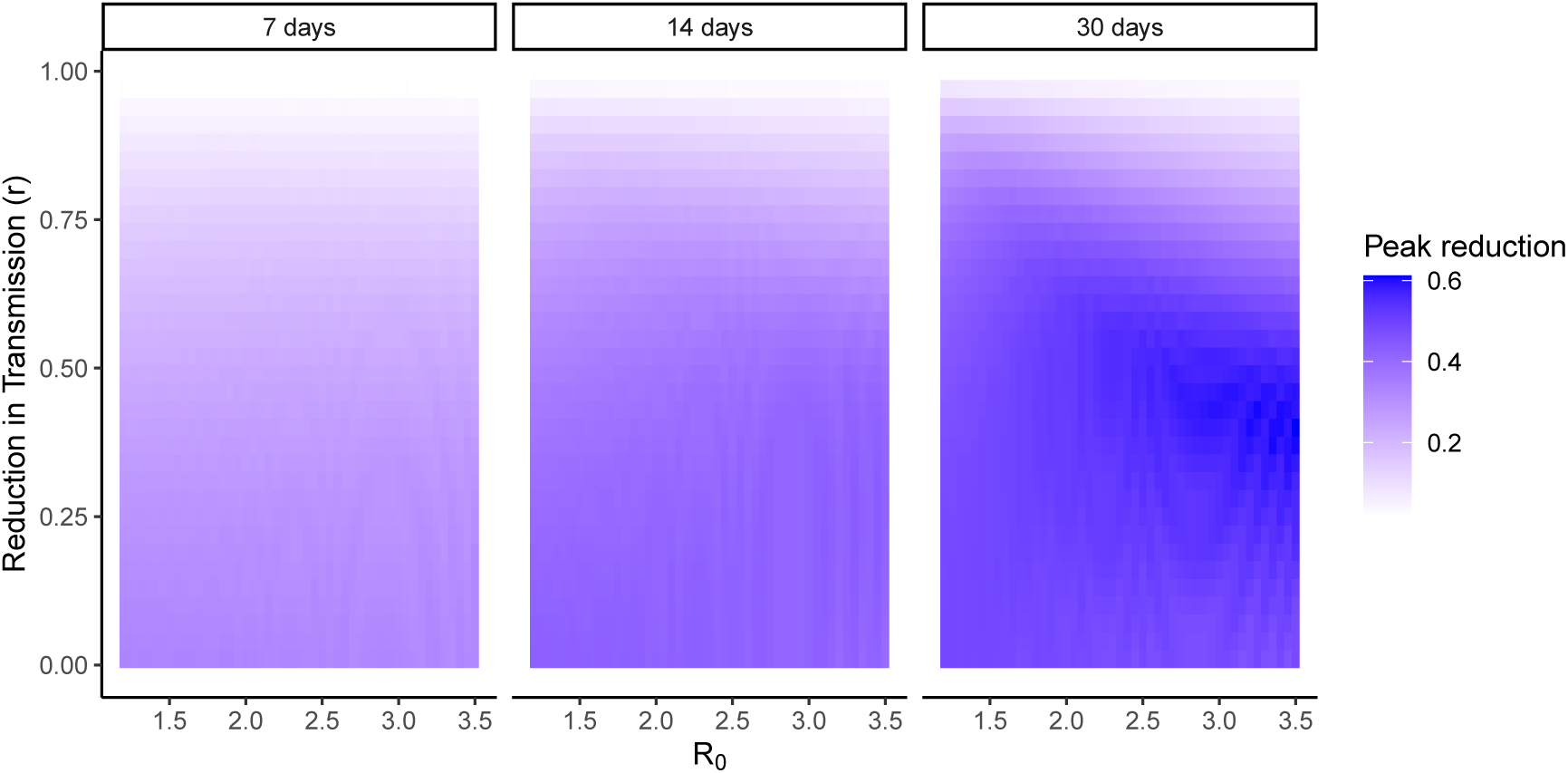
Reduction in peak time corresponding to optimal *t*_0_ to initiate social distancing as a function of *R*_0_ (x axis), reduction in transmissibility during social distancing *r* (y axis), and length of social distancing intervention (7, 14 and 30 days). For all simulations *γ* = 1*/*10.

## Appendix 4: Simulations for the optimal *t*_0_ to flatten the epidemic when an effective intervention is available in the feature

Figure (11) shows that as *t*_*I*_ decreases so does the optimal social distancing time if the goal is to flatten the epidemic curve.

**Figure 11:**
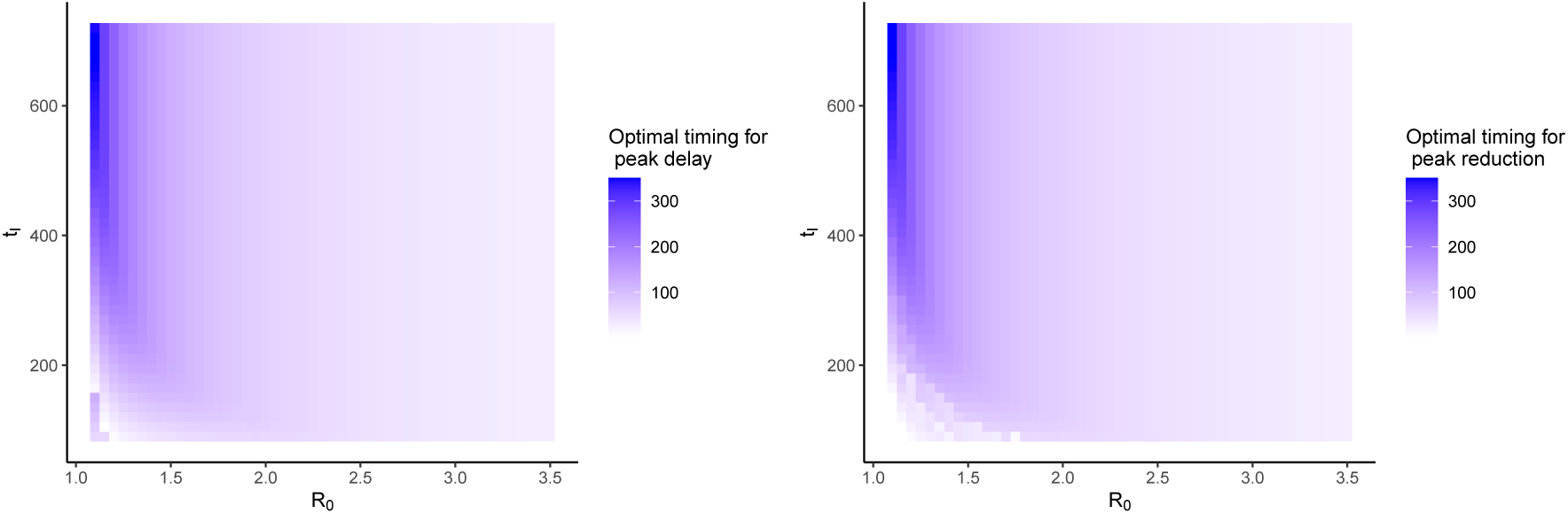
(left) Optimal *t*_0_ to initiate social distancing to maximize the delay of the epidemic peak as a function of *R*_0_ (x axis), and time to pharmacological intervention (*t*_*I*_) (y axis). (right) Optimal *t*_0_ to initiate social distancing to maximize the reduction of the epidemic peak as a function of *R*_0_ (x axis), and time to pharmacological intervention (*t*_*I*_) (y axis). With *γ* = 1*/*10, *T* = 30, *r* = 0.55, *I*(0) = 0.01%.

## Footnotes

1 Strictly speaking, it is 150 days after 0.01% of the population has been infected, as *I*(0) = 0.01%.

